# Comparing Artificial Intelligence versus Human Screening in Systematic Reviews

**DOI:** 10.64898/2026.07.01.26356995

**Authors:** Marco Gorici, Abel Torres-Espin, Mark Oremus

## Abstract

**Introduction:** Systematic reviews are essential for informing health policy and practice. Artificial intelligence (AI) automates the article screening process and produces time savings, although the performance of AI screening compared to traditional human screening remains uncertain. We undertook this study to compare the performance of two agentic AI tools, namely Loon Lens^TM^ and Catchii, to one another and to humans at the title and abstract screening level. We also compared Loon Lens to humans at the full-text screening level.

**Methods:** We developed a *de novo* research question on the association between any of three ambient air pollutants – carbon monoxide, ozone, nitrogen dioxide – and the onset or worsening of Parkinson’s disease. A health sciences librarian developed the literature search strategy and we proceded with human screening guided by PRISMA. We uploaded the retrieved citations and the eligibility criteria to both AI tools and compared screening results using sensitivity, specificity, positive predictive value, negative predictive value, concordance, kappa, and F1 score. We compared the calculated performance statistics to those obtained by naïve guessing and regressed concordance (agree or disagree with the human reference standard) onto confidence scores provided by Loon Lens, which assigned a confidence level (‘Very High’, High’, Medium’, or ‘Low’) to each of its screening decisions. Human screening was the reference standard against both AI tools; Catchii was the reference standard against Loon Lens.

**Results:** At title and abstract screening, Loon Lens outperformed Catchii when humans were the reference standard. At full-text screening, most disagreements centered around articles Loon Lens included and humans excluded. At both screening levels, higher confidence scores were associated with lower odds of disagreement between Loon Lens and human screeners.

**Discussion:** Given the panoply of available AI screening tools and their differential performance, plus the rapidly evolving nature of AI technology, researchers should pilot test their chosen tool at the start of each review. Sensitivity, kappa, and F1 are the optimal performance statistics to employ, especially at title and abstract screening, where the imbalance between proportions of included and excluded citations can inflate concordance and negative predictive value.

**What is new?:** *Key findings:*

- Strongest screening agreement seen among excluded articles
- Abstract wording affects AI tools’ performance
- Model instructions and wording of eligibility criteria also affect performance

*What this adds to what is known?:*

- Up-to-date evidence comparing the performance of two AI tools to one another and humans at title/abstract and full-text screening

*What is the implication?:*

- AI tools should be pilot tested at the outset of any review

**Plain Language Summary:** Systematic reviews help health practitioners and policy makers decide on courses of action. However, these reviews can take a long time to finish. We wanted to know whether artificial intelligence (AI) tools can replace humans when reviewing hundreds or thousands of research papers for inclusion in systematic reviews (a practice called ‘screening’). We compared two AI tools, Loon Lens™ and Catchii, to human screeners in the area of exposure to three air pollutants (carbon monoxide, ozone, or nitrogen dioxide) and occurrence of Parkinson’s disease. At title-and-abstract screening, the two AI tools had the strongest agreement with each other and with human screeners for papers that were not relevant. Loon Lens generally outperformed Catchii when both AI tools were compared to humans. At full-text screening, disagreements mainly happened when Loon Lens included a paper that humans decided to exclude. Loon Lens assigned each screening decision a ‘confidence’ level (Very High, High, Medium, Low): the higher the confidence level, the less often Loon Lens disagreed with humans. Overall, AI can serve alongside human screeners, though it is not ready to replace them yet. Researchers should test their chosen AI tool when starting a review. This will help them get the best performance out of their tool.

## 1. Introduction

### 1.1. Background

Artificial intelligence (AI), and specifically large language models (LLMs), can streamline the screening process in systematic reviews [1–3]. This process is often cited as the most time-consuming portion of any review, potentially requiring up to 20% of the total person-days needed to complete a review [4]. Although lengthy timeframes promote methodological rigor, some reviews might not be completed in time to inform health policy and practice decisions. AI tools, which speed up screening times in systematic reviews [5], can mitigate this problem if they are able to replicate human screening decisions [6]. While AI tools hold promise, evidence supporting their use is in its infancy and guidelines for employing AI in evidence synthesis are still under development [7].

### 1.2. AI in Systematic Reviews

Several studies have compared AI tools to humans in systematic review screening [6,8–11]. For example, at title and abstract (TiAb) screening, Loon Lens^TM^ (Loon Inc., Ottawa, Ontario) demonstrated 96% concordance with human screeners in eight published systematic reviews [9]. At full-text (FT) screening, Loon Lens produced 83% concordance with humans in the same pool of eight reviews [10]. Overall, the use of AI tools in screening has generally demonstrated fair-to-high concordance and low false negative rates compared to human screeners [5,6,11–13].

While these findings show promise for AI tools, most existing studies [5,6,9–11,13] have based their AI versus human comparisons on published systematic reviews. Given the large amount of public text data used in most LLM training, the possibility of test set contamination in these previous evaluations is high. This can bias comparisons in favor of the AI tools due to model memorization, limiting the relevance and generalizability of the reported findings.

### 1.3. Study Objectives

We undertook this study to compare the performance of two AI tools, Catchii [14,15] and Loon Lens, and human screeners to one another at the TiAb level, and Loon Lens to humans at the FT level. Our work is novel because we conducted the screening comparisons using a *de novo* research question developed specifically for this project.

## 2. Methods

### 2.1. Literature Search Strategy

Guided by PRISMA [16], we conducted a literature search to obtain peer-reviewed publications on the association between any of three ambient air pollutants – carbon monoxide, ozone, or nitrogen dioxide – and the onset or worsening of Parkinson’s disease. We searched PubMed and Scopus from database inception through December 12, 2025. We consulted a health sciences librarian to devise the literature search strategy (Appendix A). Table 1 contains an overview of the eligibility criteria. We excluded citations without abstracts because the AI tools were not programmed to screen these items at TiAb.

**Table 1.** Eligibility criteria

| Inclusion | Exclusion |
| --- | --- |
| Primary or secondary observational studies of a human population: | Non observational studies such as experimental (clinical trials, especially for a drug), qualitative studies, or case reports, case studies, case-series: |
| Examples of this criterion include, cross sectional, longitudinal study (cohort/panel both count), case controls, etc., | Look for key words in title and abstract that suggest an article is a review, a meta-analysis, commentary, editor, a summary, or perhaps a book chapter. |
| Prevalence/Incidence are included in relevant observational studies. | Generally, studies should not make mention of ‘patients’. This mention is indicative of case-series instead of the type of observational studies we are looking for (cross sectional, longitudinal study [cohort/panel both count], case controls, etc.) |
| Air pollutants to be mentioned: Carbon Monoxide (CO), Ozone (O3), Nitrogen Dioxide (NO2): | No air pollutant is mentioned: |
| At least one pollutant must be mentioned in TiAb or FT. | The study only mentions ‘Particulate Matter’; a separate set of particles from the air pollutants mentioned. |
| Setting can be either outdoors or indoors. | Particulate matter consists of PM, PM2.5, PM10, and is also known as ‘UFP’ or ‘Ultra fine particulate matter’. |
| Parkinson's is explicitly mentioned as an adverse health outcome: | Parkinson's is not explicitly mentioned as an adverse health outcome: |
| Parkinson's as an adverse health outcome must be treated as at least one outcome of interest and may additionally include Alzheimer's disease or similar diseases as evaluated health outcomes. | ‘Neurodegenerative’ choice of words instead of ‘Parkinsons’ may not be an appropriate exclusion criterion. |
|  | Select ‘Maybe’ option if the article uses these groupings to investigate if Parkinson's is explicit in FT so long as every other criterion is met. |
| Parkinson's is explicitly mentioned as an adverse health outcome related to air pollution: | Parkinson's is not mentioned as related to air pollutants above: |
| Parkinson's must be investigated as a health outcome related to either of the three defined air pollutants. | Studies that do not link any of the three defined air pollutants to Parkinson's disease are excluded at TiAb, except when this is unclear and every other inclusion criterion is met. |
| Fatality rates for Parkinson's count for the inclusion criteria. |  |
| Parkinsons hospitalizations or hospitalizations because of individuals with Parkinson's exposed to air pollution can be included. |  |
| The article must be written in English. | The article is not written in English. |

### 2.2. Human and AI Screening Processes

Following duplicate removal, we imported citations into Covidence (Covidence, Melbourne, Australia) and each was screened independently by two humans at TiAb. Citations were promoted to FT if each inclusion criterion was met, or the combination of responses to all the inclusion criteria was a mix of met or unsure. ‘Unsure’ meant the criterion could not be assessed based on the information presented in the title or abstract. At FT, the unsure option was no longer available; screeners included articles if all the eligibility criteria were met. Disagreements between screeners were addressed by consensus or a third screener.

Catchii and Loon Lens are hosted on secure servers. We accessed the tools through registered accounts and a browser interface. At TiAb, both tools required us to paste the eligibility criteria, in point form, into prespecified open-text fields. The next step was to upload the deduplicated list of retrieved citations from the literature search [9,10,14]. Both AI tools produced spreadsheets identifying the citations they flagged for inclusion.

The articles that human screeners advanced to FT from TiAb served as the basis of the FT comparison. We uploaded the eligibility criteria and PDFs of these articles onto the Loon Lens platform. Catchii was excluded from the FT comparison because it did not offer a FT screening service.

### 2.3. Analysis

Catchii and Loon Lens were compared to humans as the reference standard at TiAb; Loons Lens was also compared to Catchii as the reference standard at TiAb. We compared Loon Lens to humans as the reference standard at FT. We created a 2 x 2 confusion table for each comparison and generated seven performance statistics from the information in each table: sensitivity, specificity, positive predictive value (PPV), negative predictive value (NPV), concordance, kappa, and F1. We estimated 95% confidence intervals (CIs) for the statistics using the percentile bootstrap method with 1,000 resamples and graphed the point estimates for the first five statistics on radar charts.

For each article screened at TiAb and FT, Loon Lens provided a confidence score (‘very high’, ‘high’, ‘medium’, or ‘low’) to describe the degree of certainty in its include/exclude decisions. To explore whether higher confidence scores were associated with lower odds of disagreement in screening decisions between Loon Lens and human screeners, we regressed ‘concordance’ (proportion of articles for which Loon Lens’ include or exclude decisions agreed with human screeners) on the confidence scores. In separate simple logistic regression models for the TiAb and FT screening levels, we operationalized concordance as a two-level variable: ‘agree’ (reference category) flagged articles where Loon Lens and humans produced the same decision; ‘disagree’ identified articles where the decision differed. For the TiAb analysis, the ‘medium’ confidence score was used as the reference category (relabeled ‘medium/low’) because Loon Lens assigned a ‘low’ confidence score to only one article, which was grouped with the ‘medium’ confidence articles. We did not conduct this analysis for the Catchii versus human comparison because Catchii did not provide confidence scores.

To compare the AI tools’ performance to non-informative or naïve guessing, we assumed all the screened articles should be excluded at TiAb or FT (majority class guessing), and the articles included by the reference standard were done so erroneously. We constructed confusion tables and calculated performance statistics based on these assumptions. The resulting performance statistics represented the findings one would obtain through guessing based on the majority class as the null hypothesis. We tested whether the actual performance of the AI tools differed from naïve guessing by verifying whether the lower bound of each performance statistic’s percentile bootstrapped 95% CI exceeded the corresponding point estimate for a naïve result. If so, then we concluded the AI tool performed better than naïvely guessing based on the majority class of exclusion for the statistic in question.

We utilized R v4.5.1 (The R Foundation for Statistical Computing, Vienna, Austria) and multiple R packages in the analysis [17–30]. The R code and screening results are available on GitHub [31]. This study did not require ethics approval because it was based on published articles in the scientific literature.

## 3. Results

### 3.1. Article Flow through Screening

After removing duplicates, 793 articles from PubMed and Scopus were screened at TiAb: human screeners sent 61 articles to FT, Catchii sent 58, and Loon Lens sent 44. Of the 61 articles sent by human screeners to FT, Loon Lens included 52 articles and humans included 41. Figures 1 and 2 present the flow of articles through the screening process. Confusion and concordance tables are shown in Appendix B. Radar charts are shown in Appendix C.

**Figure 1.**
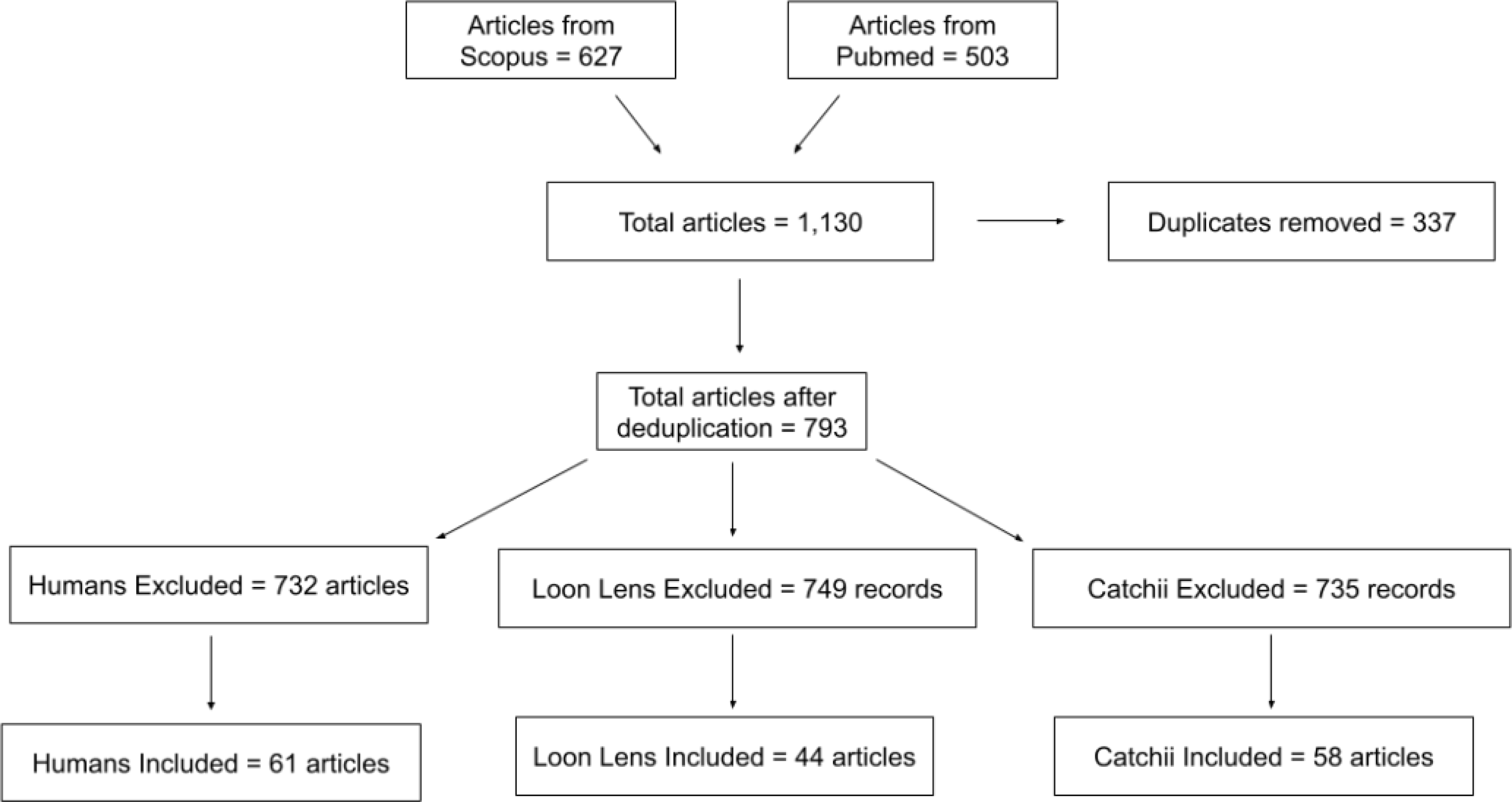
Article flow through title and abstract screening

**Figure 2.**
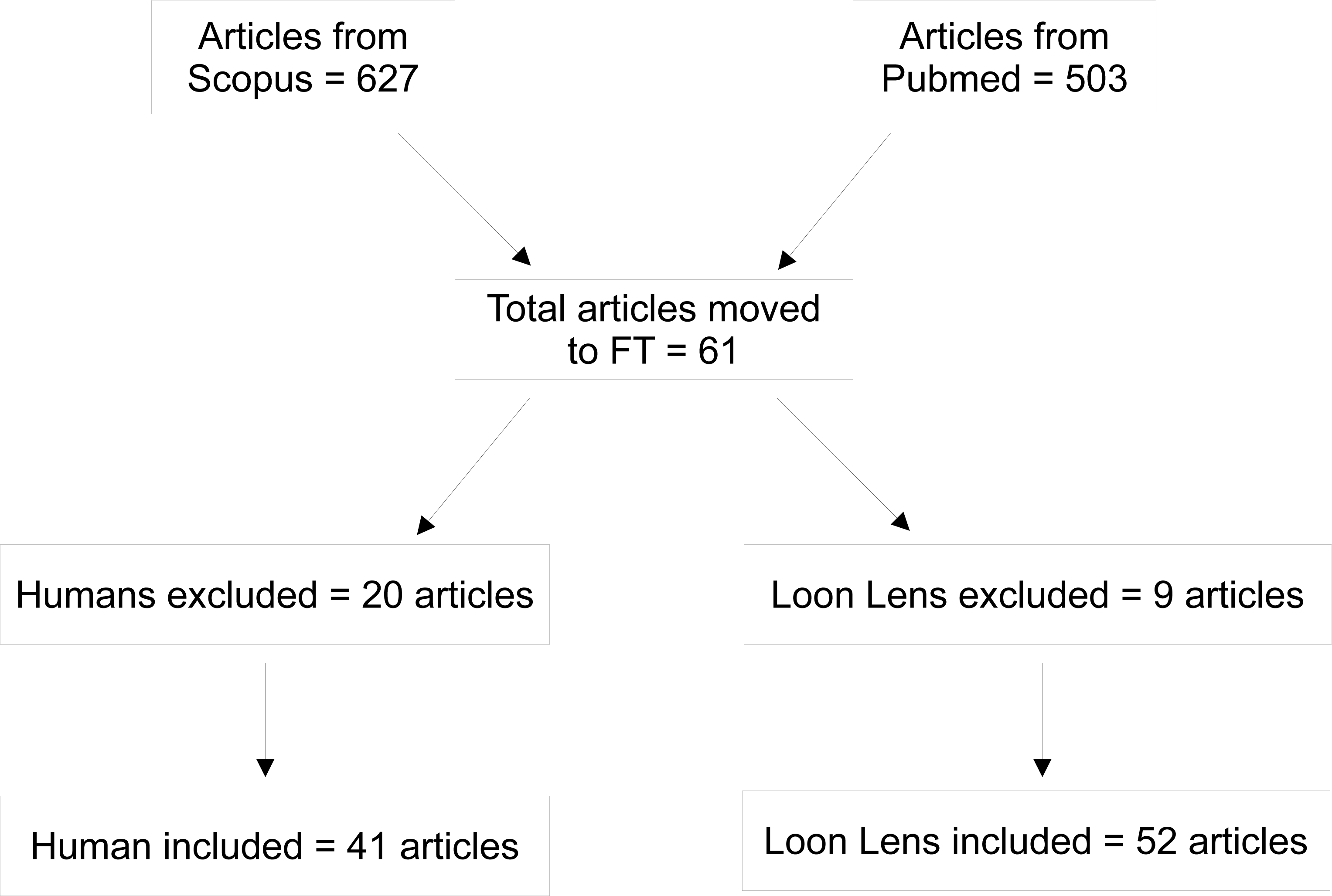
Article flow through full-text screening FT = full-text screening.

### 3.2. Title and Abstract Screening

#### 3.2.1. Catchii versus Humans

Catchii correctly classified 30 out of 61 articles as included and 704 out of 793 as excluded (Appendix B – Table B.1). Figure 3A displays the performance statistics. In total, Catchii included 7% (n = 58) of the 793 screened articles with 93% (n = 734) of the screening decisions in concordance with human screeners.

**Figure 3.**
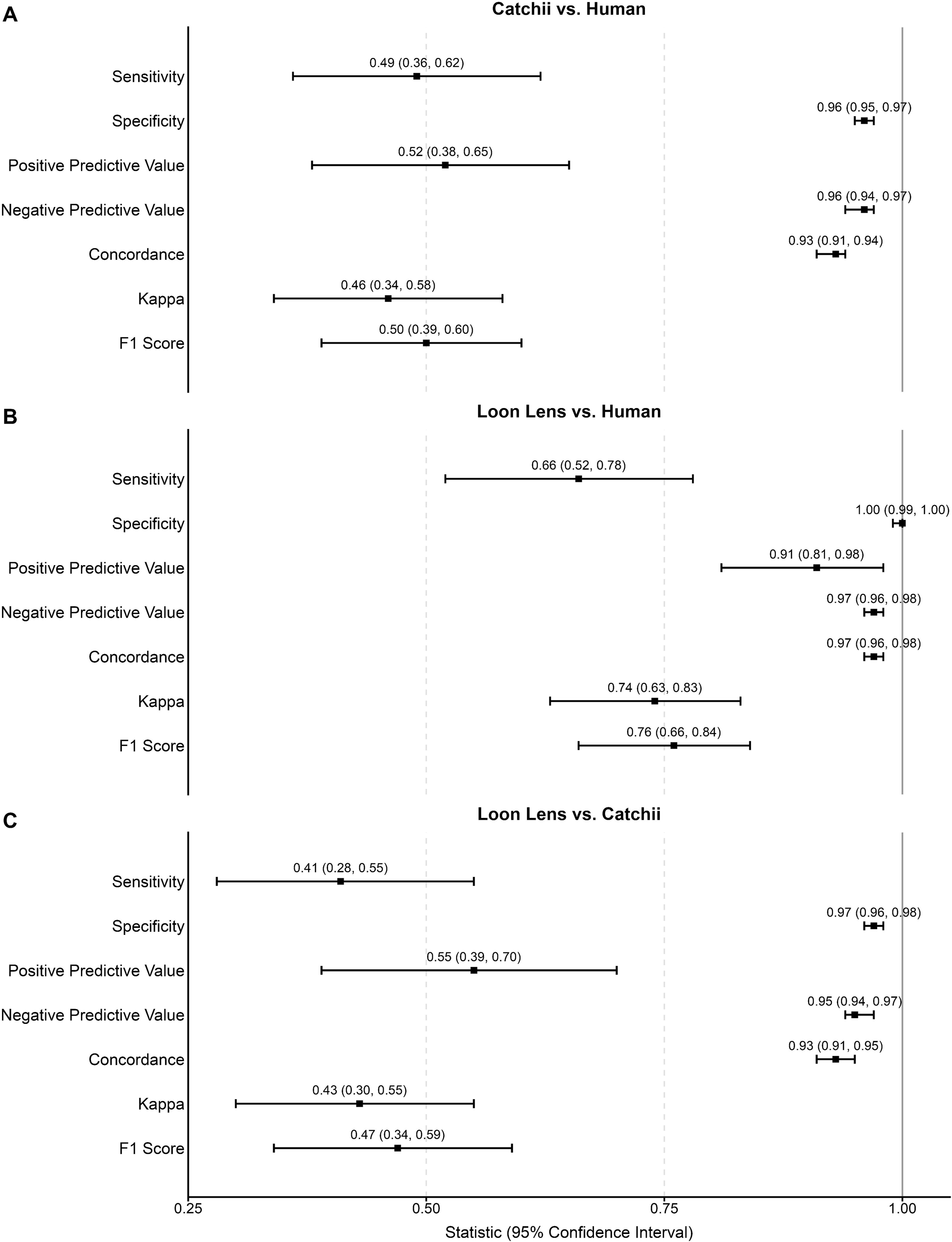
Performance statistics – title and abstract screening

#### 3.2.2. Loon Lens versus Humans

Loon Lens included 40 of the 61 articles humans included, and excluded 728 of the 732 articles humans excluded (Appendix B – Table B.2). Figure 3B shows the performance statistics. Overall, Loon Lens included 6% (n = 44) of the 793 screened articles and attained roughly 97% (n = 768) concordance with human screeners.

##### 3.2.2.1 Confidence Scores and Concordance

Loon Lens and humans disagreed on the screening results of 25 articles at TiAb (Appendix B –Table B.3). The largest number of disagreements (n = 11) occurred at the ‘medium’ confidence score. When regressing concordance on confidence scores, the odds of disagreement were lower for the high and very high confidence scores, compared to the medium/low confidence score (Table 2).

**Table 2.**
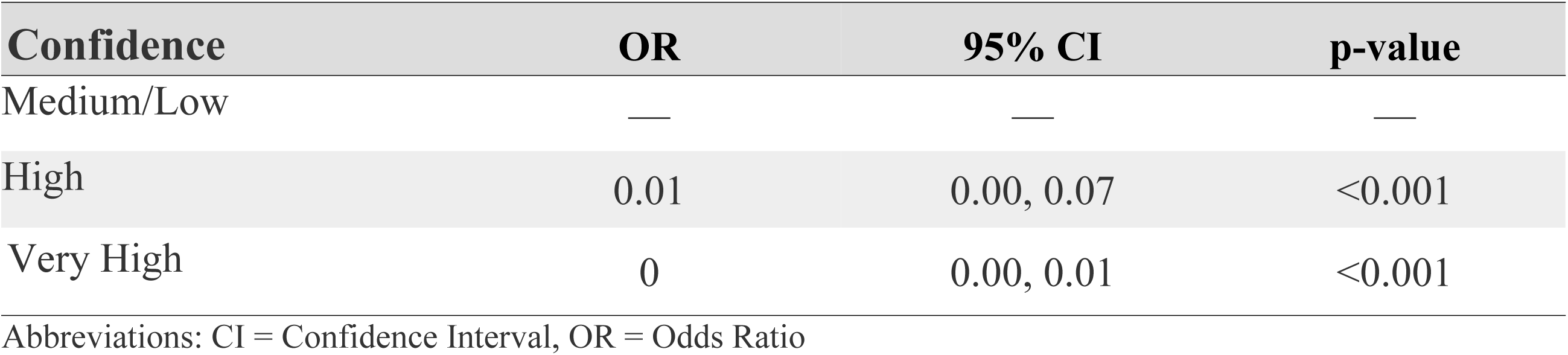
Association between Loon Lens screening confidence and disagreement between artificial intelligence and human screening – title and abstract screening

#### 3.2.3. Loon Lens versus Catchii

At TiAb, with Catchii as the reference standard, Loon Lens included 24 of the 58 articles that Catchii included, and excluded 715 of the 735 articles that Catchii excluded (Appendix B – Table B.4). Figure 3B displays the performance statistics. Loon Lens included an additional 20 articles and excluded an additional 34 articles, compared to Catchii. Concordance between the AI tools was 93% (n = 739).

### 3.3. Full-Text Screening

Loon Lens included 40 of the 41 articles humans included at FT, and excluded 8 of the 20 articles humans excluded (Appendix B – Table B.5). Performance statistics were sensitivity of 98% (95% CI: 92%, 100%), specificity of 40% (95% CI: 19%, 64%), PPV of 77% (95% CI: 65%, 88%), and NPV of 89% (95% CI: 60%, 100%). Concordance was 79% (n = 48), F1 was 86% (95% CI: 78%, 93%), and kappa was 0.44 (95% CI: 0.17, 0.65).

#### 3.3.1. Confidence Scores and Concordance

Loon Lens and humans disagreed on the screening results of 13 articles at FT (Appendix B – Table B.6). Almost half (n = 6) of the disagreements occurred at the ‘low’ confidence score. When regressing concordance on the confidence scores, the odds of disagreement were lowest for the very high versus low confidence scores (Table 3). Although the odds were also lower for the high and medium confidence scores, these results were not statistically significant.

**Table 3.** Association between Loon Lens screening confidence and disagreement between artificial intelligence and human screening – full-text screening

| Confidence | OR | 95% CI | p-value |
| --- | --- | --- | --- |
| Low | — | — | — |
| Medium | 0.75 | 0.07, 8.16 | 0.8 |
| High | 0.50 | 0.04, 5.94 | 0.6 |
| Very High | 0.02 | 0.00, 0.15 | <0.001 |

Abbreviations: CI = Confidence Interval, OR = Odds Ratio

### 3.4. Comparison of Catchii and Loon Lens Performance to Naïve Guessing

Assuming all screened articles should be excluded (naïve guessing), chance led to the inclusion of 61 articles at TiAb by the human reference standard, 58 articles at TiAb by the Catchii reference standard, and 41 articles at FT by the human reference standard. Thus, at TiAb, concordance achieved by chance alone was 92% for Catchii and Loon Lens versus humans, and 93% for Loon Lens versus Catchii. Concordance was 67% for Loon Lens versus humans at FT. Appendix D – Table D.1 and Figure D.1 show all the performance statistics and confusion tables obtained by chance alone.

Across all four comparisons, Loon Lens and Catchii performed better than naïve guessing on sensitivity, NPV, kappa, and F1 (Appendix D – Table D.2). They did not perform better on specificity because the latter will always attain values of 100% under naïve guessing, given that all articles should be excluded (Appendix D – Figure D.2). PPV was undefined under naïve guessing because the cells of the confusion tables used to calculate it were 0 (Appendix D – Figure D.2). Results for concordance showed Loon Lens performed better than naïve guessing against humans at TiAb and FT, though not against Catchii at TiAb. Catchii’s concordance was no better than naïve guessing against humans at TiAb.

## 4. Discussion

Compared to humans at TiAb, Loon Lens correctly included many relevant articles and excluded almost all irrelevant articles, whereas Catchii included fewer relevant articles and excluded nearly all irrelevant articles. For the Loon Lens versus Catchii comparison at TiAb, the low sensitivity indicated disagreement on which articles to include. When comparing Loon Lens to human screeners at FT, the AI tool performed best at including articles that were included by humans. The Loon Lens to human comparisons were fairly consistent with previous studies, where Loon Lens’ performance across all metrics ranged from 80% to 100% [9,10].

Given the comparison of estimated versus naïve guessing performance statistics, little evaluative weight should be placed on specificity and PPV. Concordance is also a questionable statistic to use when evaluating AI tools at TiAb because the small proportions of included citations at this screening level (∼ 7% to 8% in our case) will invariably produce high estimates of concordance by chance or naïve guessing alone. Similarly, the large number of citations that are typically excluded at TiAb will produce high estimates of NPV.

Sensitivity, kappa, and F1 are therefore likely to be the most useful statistics to utilize when comparing AI tools to one another or humans in systematic review screening. In our study, Loon Lens and Catchii performed better than naïve guessing on these three statistics across all comparisons. At TiAb, Loon Lens outperformed Catchii on sensitivity, kappa, and F1 when humans were the reference standard.

Loon Lens’ confidence scores enabled us to assess reasons for disagreement with human screeners. In one case that exemplified many disagreements, Loon Lens excluded an article [32] with very high confidence at TiAb that humans advanced to FT. Humans promoted the article because its title and abstract provided insufficient information to assess its relevance. Human screeners grapple with ambiguity at TiAb by promoting articles to FT for further scrutiny. Loon Lens appeared to make binary include/exclude decisions based on the presence or absence of key words or phrases.

We suspect high-level abstracts and inconsistencies between abstracts and full reports [33,34] will generate most disagreements between AI tools and humans at TiAb. As such, AI tools will be more likely to exclude potentially relevant articles at TiAb. AI tools might perform better if researchers first provide instructions to LLMs on how to behave when ambiguity is present, e.g., promote ambiguous articles to FT.

At FT, the same article [32] discussed above was included with low confidence by Loon Lens and excluded by humans. Loon Lens included the article for containing the word ‘parkinsonism’ and listing symptoms commonly linked to Parkinson’s disease. Human screeners excluded the article because the authors did not examine diagnosed Parkinson’s disease as an outcome, although the article required a thorough read to make this determination. AI tools might erroneously include ambiguous articles that contain lists of words seen repeatedly in previously included articles. While explicit model instructions can mitigate some over inclusion, crafting these instructions can pose a challenge. For example, truly relevant articles in our case often looked at ‘parkinsonism or Parkinson’s disease’, which prevented us from instructing the AI tools to exclude articles containing the word ‘parkinsonism’.

When analyzing disagreements between Loon Lens and Catchii, we discovered Catchii excluded articles whose titles or abstracts only employed abbreviations for the air pollutants of interest (“CO”, “NO_2_”, or “O_3_”). Conversely, Loon Lens promoted these articles to FT, even when the full names of the pollutants were absent from titles or abstracts. Our eligibility criteria did not mention the abbreviations. Lexical ambiguities such as abbreviations or short forms of words can be problematic for LLMs, especially if a general-purpose model is used instead of a domain-specific one, such as PubMedBERT [35] in health. Since AI tools vary in how they handle ambiguity, depending on the LLMs used and knowledge systems built around them, we recommend researchers explicitly provide important abbreviations or short-form words in the eligibility criteria.

The rationales that Loon Lens and Catchii provided for their inclusion/exclusion decisions, which are shown in the .csv files on GitHub [31], may serve as the basis for pilot testing AI tools prior to the launch of actual screening. Similar to a calibration pilot for human screeners, researchers can give their chosen AI tool a random sample of citations to screen, with humans verifying the AI results and rationales, and refining the instructional prompts and eligibility criteria accordingly. Pilot-testing AI tools maps onto the ‘human-in-the-loop’ workflow recommended when integrating AI into systematic reviews [36]. See Appendix E – Figure E.1 for a suggested ‘human-in-the-loop’ workflow for TiAb and FT screening.

### 4.1. Strengths and Limitations

This study’s major strength was the use of a *de novo* research question to prevent test-training contamination. The common practice of employing published systematic reviews in comparative evaluations of AI versus human screeners can lead to inflated AI performance, as the models underlying the AI tools may have already been trained on the results of the published reviews. A limitation of the study was the inability to assess trade-offs between performance and efficiency because we did not collect screening times for the AI tools and humans.

### 4.2. Recommendations

The metaphor of the ‘black box’ is commonly used to highlight the risk of interacting with AI models whose internal workings are not transparent [37–39]. The absence of transparency underscores the need to choose models with rigorous and publicly available evaluations of performance. Additionally, as with traditional human screening, AI tools should be pilot tested on a subset of articles prior to the commencement of the full screening process, and protocols revised accordingly. This recommendation is especially important for tools like Loon Lens and Catchii, which can operate by zero-shot inference. Since multiple screening runs can yield different results, the pilot testing phase should be repeated to assess test-retest reliability, although the literature does not specify a minimum number of runs. AI tools can also be paired with independent human screeners to save time, with conflicts assessed by a third-party human.

To promote transparency, we encourage the developers of AI tools to follow the example of Loon Lens and evaluate tool performance [9,10]. Another example of transparency is ASReview, which provides documentation on its website [40] and open-access repositories on GitHub [41]. At a minimum, we encourage the developers of AI screening tools to provide their codebases, underlying LLMs, knowledge systems, and instructional prompts.

### 4.3. Conclusion

This study investigated how AI tools, specifically Loon Lens and Catchii, compared to humans in systematic review screening. Through a *de novo* research question on the association between carbon monoxide, ozone, nitrogen dioxide, and Parkinson’s disease, we found that Loon Lens outperformed Catchii in comparison to the reference standard of human screeners. Our work contributes to the evidence base for using AI tools in systematic reviews. Future research can incorporate time and cost metrics into the comparative evaluation process, explore the effect of word choice in specifying eligibility criteria, and assess additional AI screening tools.

## Data Availability

The R code and AI screening results are available on GitHub: https://github.com/moremus/AI_versus_Human_Screening.git

## CRediT authorship contribution statement

**Marco Gorici –** Conceptualization, Data Curation, Formal Analysis, Investigation, Software, Visualization, Writing – original draft. **Abel Torres-Espin** – Methodology, Validation, Writing – review & editing. **Mark Oremus** – Conceptualization, Data Curation, Formal Analysis, Methodology, Project Administration, Resources, Software, Supervision, Validation, Writing – review & editing.

## Declaration of competing interests

The authors declare no conflicts of interest.

## Acknowledgements

We thank Ashton Albertie, Alexandru Gagu, Ella Mironescu, and Noah Ibanez for serving as screeners, and Jackie Stapleton for developing the literature search strategy. We also thank Ghayath Janoudi, Founding CEO/CTO of Loon Inc., for providing access to Loon Lens^TM^. Loon Inc. had no say in the design and conduct of the study, nor in the interpretation or presentation of results.

## Funding

This research did not receive any specific grant from funding agencies in the public, commercial, or not-for-profit sectors.

## Ethical approval

This study was exempted from ethics approval because it relied solely on publicly-available, published data.

## Declaration of generative AI in the manuscript preparation process

During the preparation of this work, the authors used DALL·E 3 (via ChatGPT) to create Appendix E – Figure E.1 and Claude Sonnet 4.6 (Anthropic, claude.ai) to create the graphical abstract. After using these tools, the authors reviewed and edited the content as needed and take full responsibility for the content.

## Notes

### Competing Interest Statement

The authors have declared no competing interest.

