## Supplemental Files for "Comparing Artificial Intelligence versus Human Screening in Systematic Reviews"

**Appendices**

**Appendix A. Search Criteria**

Pubmed: (“air pollut*” OR “particulate matter” OR “Air Pollutants”[Mesh] OR “Air Pollution"[Mesh] OR “carbon monoxide” OR “carbon monoxide”[Mesh] OR “nitrogen dioxide” OR “NO2” OR “ozone” OR “O3”) AND (“parkinson*”).

Scopus: (“carbon monoxide” OR “nitrogen dioxide” OR “NO_2_” OR “ozone” OR “O_3_”) AND (“parkinson*”).

**Appendix B. Confusion and Concordance Tables**

| **Table B.1.** Confusion Table - Title and Abstract Screening:  Catchii versus Human Screening | | | |
| --- | --- | --- | --- |
|  |  | Human Screening* | |
|  |  | Include | Exclude |
| Catchii | Include | 30 | 28 |
|  | Exclude | 31 | 704 |
| *Reference standard | | | |

| **Table B.2.** Confusion Table - Title and Abstract Screening: Loon Lens versus Human Screening | | | |
| --- | --- | --- | --- |
|  |  | Human Screening* | |
|  |  | Include | Exclude |
| Loon Lens | Include | 40 | 4 |
|  | Exclude | 21 | 728 |
| *Reference standard | | | |

| **Table B.3.** Loon Lens Confidence Level by Screening Concordance - Title and Abstract Screening | | |
| --- | --- | --- |
| Concordance | | |
|  | Agree | Disagree |
|  | (n = 768) | (n = 25) |
| Confidence |  |  |
| Very High | 687 (89.5%) | 6 (24.0%) |
| High | 79 (10.3%) | 7 (28.0%) |
| Medium | 2 (1.0%) | 11 (44.0%) |
| Low | 0 (0%) | 1 (4.0%) |

**Chi-square test:** p < 0.0001

| **Table B.4.** Confusion Table - Title and Abstract Screening: Loon Lens versus Catchii | | | |
| --- | --- | --- | --- |
|  |  | Catchii* | |
|  |  | Include | Exclude |
| Loon Lens | Include | 24 | 20 |
|  | Exclude | 34 | 715 |
| *Reference standard |  |  |  |

| **Table B.5.** Confusion Table - Full Text Screening: Loon Lens versus Human Screening | | | |
| --- | --- | --- | --- |
|  |  | Human Screening* | |
|  |  | Include | Exclude |
| Loon Lens | Include | 40 | 12 |
|  | Exclude | 1 | 8 |
| *Reference standard | | | |

| **Table B.6.** Loon Lens Confidence Level by Screening Concordance - Full-text Screening | | |
| --- | --- | --- |
|  | Concordance | |
|  | Agree  (n = 48) | Disagree  (n = 13) |
| Confidence |  |  |
| Very High | 41 (85.4%) | 2 (15.4%) |
| High | 2 (4.2%) | 2 (15.4%) |
| Medium | 2 (4.2%) | 3 (23.1%) |
| Low | 3 (6.3%) | 6 (46.2%) |

**Chi-square test:** p < 0.0001

**Appendix C. Radar Charts Summarizing Screening Performance Metrics**

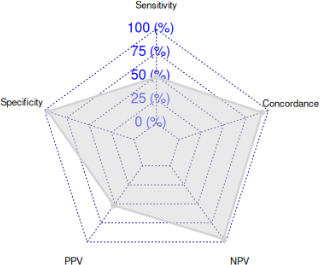

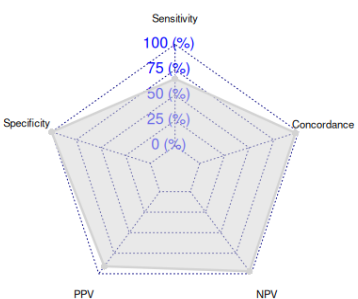

**Figure C.1. Catchii versus Human Screening (title and abstract)	Figure C.2. Loon Lens versus Human Screening (title and abstract)**

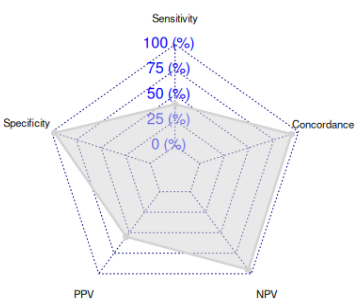

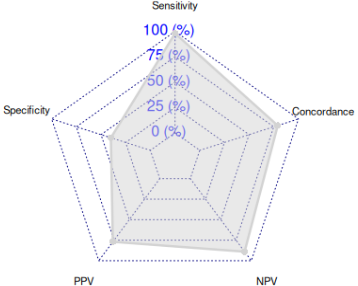

**Figure C.3. Loon Lens versus Catchii Screening (title and abstract) Figure C.4. Loon Lens versus Human Screening (full text)**

NPV = negative predictive value; PPV = positive predictive value.

**Appendix D. Class-Imbalance-Adjusted Performance Analysis**

| **Table D.1.** Performance statistics obtained by chance alone | | | | |
| --- | --- | --- | --- | --- |
|  | Title and Abstract  Screening | | | Full-text Screening |
|  | Catchi  versus  Humans | Loon Lens  versus  Humans | Loon Lens  versus  Catchii | Loon Lens  versus  Humans |
| Sensitivity | 0.00 | 0.00 | 0.00 | 0.00 |
| Specificity | 1.00 | 1.00 | 1.00 | 1.00 |
| Positive Predictive Value | Undefined | Undefined | Undefined | Undefined |
| Negative Predictive Value | 0.92 | 0.92 | 0.93 | 0.34 |
| Concordance | 0.92 | 0.92 | 0.93 | 0.34 |
| Kappa | 0.00 | 0.00 | 0.00 | 0.00 |
| F1 | 0.00 | 0.00 | 0.00 | 0.00 |

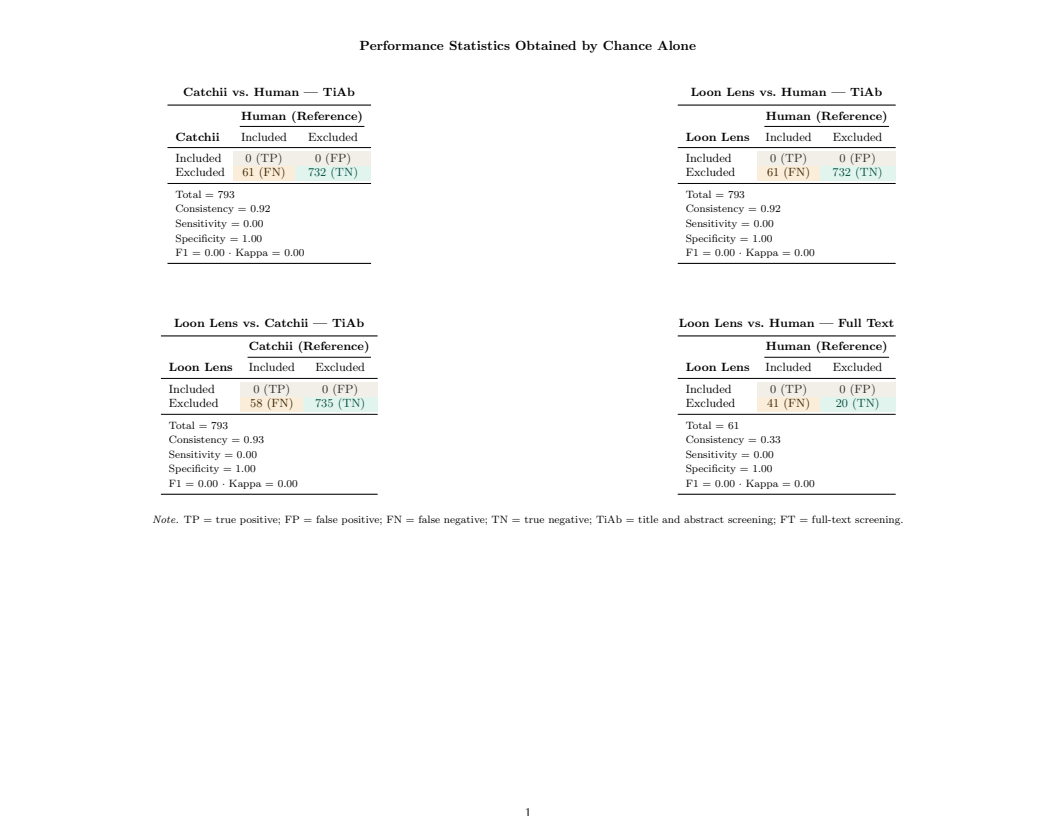
**Figure D.1. Confusion Tables Supporting Table D.1 above**

| **Table D.2.** Artificial intelligence performance statistics compared to chance alone | | | | |
| --- | --- | --- | --- | --- |
|  | Statistic | Point Estimate | Chance Alone | Exceeded* |
| Catchii vs. Human (TiAb) | Sensitivity | 0.49 | 0.00 | YES |
| Catchii vs. Human (TiAb) | Specificity | 0.96 | 1.00 | NO |
| Catchii vs. Human (TiAb) | PPV | 0.52 | Undefined | N/A |
| Catchii vs. Human (TiAb) | NPV | 0.96 | 0.92 | YES |
| Catchii vs. Human (TiAb) | Kappa | 0.46 | 0.00 | YES |
| Catchii vs. Human (TiAb) | Concordance | 0.93 | 0.92 | NO |
| Catchii vs. Human (TiAb) | F1 Score | 0.50 | 0.00 | YES |
| Loon Lens vs. Human (TiAb) | Sensitivity | 0.66 | 0.00 | YES |
| Loon Lens vs. Human (TiAb) | Specificity | 1.00 | 1.00 | NO |
| Loon Lens vs. Human (TiAb) | PPV | 0.91 | Undefined | N/A |
| Loon Lens vs. Human (TiAb) | NPV | 0.97 | 0.92 | YES |
| Loon Lens vs. Human (TiAb) | Kappa | 0.74 | 0.00 | YES |
| Loon Lens vs. Human (TiAb) | Concordance | 0.97 | 0.92 | YES |
| Loon Lens vs. Human (TiAb) | F1 Score | 0.76 | 0.00 | YES |
| Loon Lens vs. Catchii (TiAb) | Sensitivity | 0.41 | 0.00 | YES |
| Loon Lens vs. Catchii (TiAb) | Specificity | 0.97 | 1.00 | NO |
| Loon Lens vs. Catchii (TiAb) | PPV | 0.55 | Undefined | N/A |
| Loon Lens vs. Catchii (TiAb) | NPV | 0.95 | 0.93 | YES |
| Loon Lens vs. Catchii (TiAb) | Kappa | 0.43 | 0.00 | YES |
| Loon Lens vs. Catchii (TiAb) | Concordance | 0.93 | 0.93 | NO |
| Loon Lens vs. Catchii (TiAb) | F1 Score | 0.47 | 0.00 | YES |
| Loon Lens vs. Human (FT) | Sensitivity | 0.98 | 0.00 | YES |
| Loon Lens vs. Human (FT) | Specificity | 0.40 | 1.00 | NO |
| Loon Lens vs. Human (FT) | PPV | 0.77 | Undefined | N/A |
| Loon Lens vs. Human (FT) | NPV | 0.89 | 0.34 | YES |
| Loon Lens vs. Human (FT) | Kappa | 0.44 | 0.00 | YES |
| Loon Lens vs. Human (FT) | Concordance | 0.79 | 0.34 | YES |
| Loon Lens vs. Human (FT) | F1 Score | 0.86 | 0.00 | YES |
| *‘Yes’ indicates the lower bound of the 95% bootstrap confidence interval for the point estimate (Figure 3) exceeded the chance alone value; ‘No’ indicates otherwise. | | | | |
| Abbreviations: CI = confidence interval; FT = full text screening, N/A = cannot be evaluated because PPV is undefined for null, NPV = negative predictive value, PPV = positive predictive value, TiAb = title and abstract screening | | | | |

**Appendix E. Human-in-the-Loop Approach**

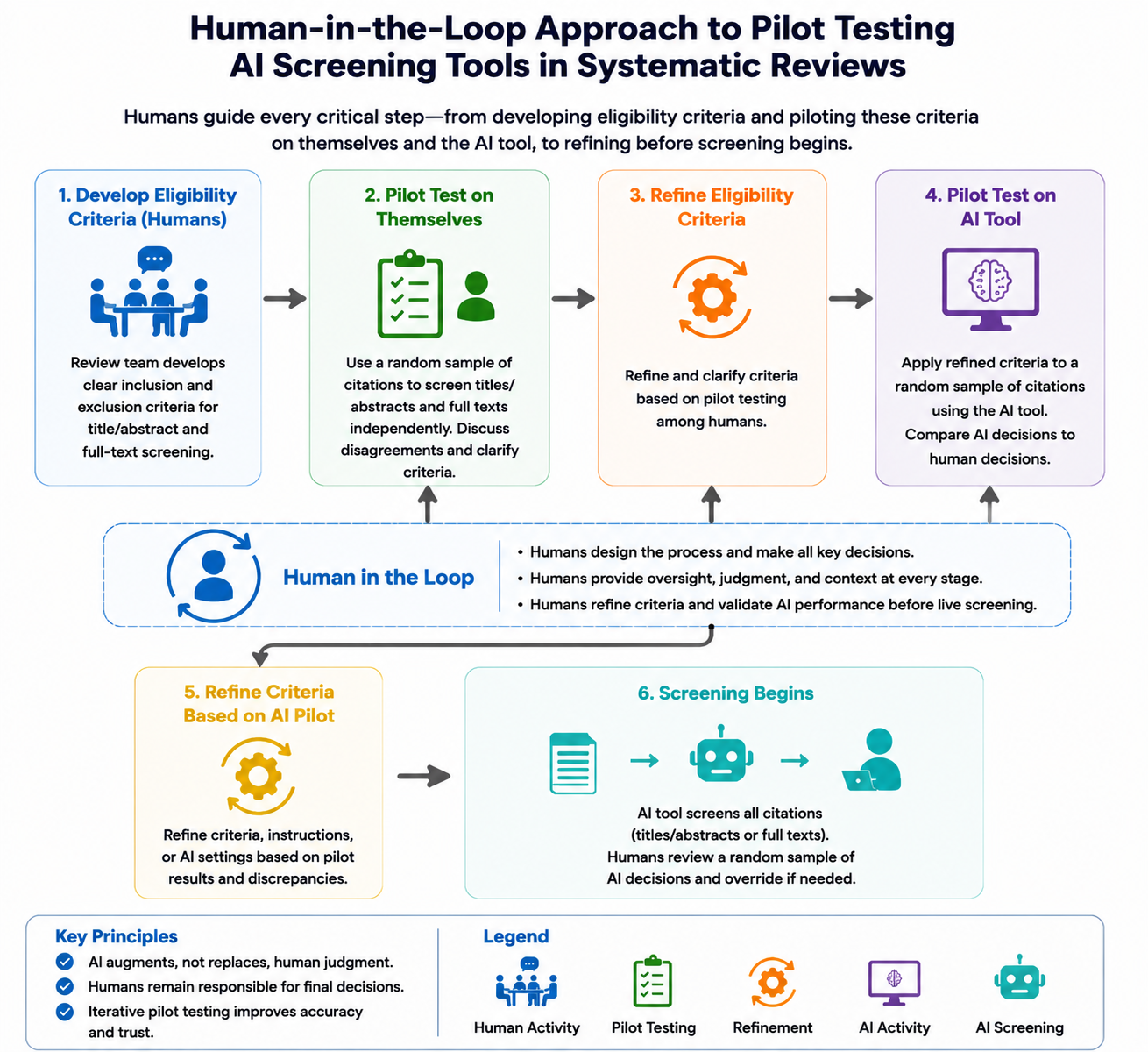
**Figure E.1. Human-in-the-Loop Approach to Pilot Testing Artificial Intelligence (AI) Tools in Systematic Reviews**

Note: The authors used DALL·E 3 (via ChatGPT) to create Figure E.1. After using this tool, the authors reviewed and edited the figure’s content as needed and take full responsibility for the figure’s content.
